# Early Cardiometabolic Vulnerability: Exploring Sleep, Depression, and Hypertension in Rural Communities

**DOI:** 10.64898/2026.01.14.26343923

**Authors:** Christopher M. Smith, Carolyn E. Horne, Jeanette M. Bennett, Brittany Butts

**Author notes:** Corresponding Author: Christopher M. Smith, School of Nursing, University of North Carolina at Charlotte, 8844 Craver Rd, Charlottee, NC, 28228.

## Abstract

**Background:** Cardiometabolic disease risk is disproportionately high in rural U.S. communities, where behavioral and metabolic determinants often intersect. However, relationships between sleep, psychological distress, and blood pressure variability remain understudied in rural communities.

**Objective:** To assess how sleep, depressive symptoms, and metabolic indicators influence blood pressure outcomes in rural adults.

**Methods:** In this exploratory descriptive cross-sectional study, survey and clinical measurement data were collected from *N* = 68 participants. Data were analyzed using descriptive statistics, bivariate correlations, and hierarchical regression.

**Results:** Age, sex, and sleep were positively associated with blood glucose. Depressive symptoms were inversely related to blood glucose and BMI. Depressive symptoms and sleep dysfunction were positively associated with blood pressures, but in some models were attenuated by sleep, whereas sleep was related to elevated blood glucose. Age was inversely associated with blood pressure and antihypertensive medication use did not account for this relationship. Among non-medicated participants, mean blood pressures were significantly elevated. In subgroup analyses, the youngest non-medicated adults (ages 29–33) had markedly higher mean arterial pressure.

**Conclusions:** Sleep-related impairment and depressive symptoms were related to BP indices and indicators of metabolic disruption and inflexibility in this rural community sample. The effect of depressive symptoms on BP outcomes was consistent with autonomic and vascular tone mechanisms but was attenuated by sleep. Younger participants exhibited disproportionately high untreated blood pressures, highlighting the possibility that cardiometabolic vulnerability may manifest earlier in rural communities than typically recognized. Results support early detection initiatives and future biobehavioral-informed research targeting younger populations.

## Introduction

Cardiometabolic disease remains a leading cause of morbidity and mortality globally, particularly in rural communities where access to care and preventive screening are limited^1^. Although lifestyle factors such as nutrition and physical activity are widely recognized pillars of cardiometabolic health,^2^ behavioral sleep and psychological health has received comparatively less attention despite evidence that poor sleep and depression exerts profound effects on both cardiovascular and metabolic systems.^3^ More specifically, sleep dysfunction and depression has emerged as critical yet modifiable risk factors in the pathophysiology of cardiometabolic disease, influencing glucose metabolism, inflammatory processes that underlie vascular dysfunction, and blood pressure (BP) regulation.^3^

## Background

Sleep and psychological health influence cardiometabolic risk through interconnected biological and behavioral pathways that operate well before clinical disease becomes apparent. Understanding these upstream mechanisms is essential for interpreting how subjective symptoms such as sleep-related impairment and depressive symptoms may translate into measurable metabolic and vascular changes. The following sections summarize current evidence describing the neuroendocrine, inflammatory, and autonomic processes linking sleep and depression to blood pressure regulation and metabolic dysfunction, with particular relevance for early and under-recognized cardiometabolic vulnerability.

Sleep disruption activates neuroendocrine and inflammatory pathways that can elevate BP and impair insulin sensitivity.^4^ Short or fragmented sleep increases sympathetic nervous system activity and hypothalamic-pituitary-adrenal (HPA) axis activation, leading to elevations in cortisol, catecholamines, and circulating inflammatory cytokines such as interleukin-6 (IL-6), C-reactive protein (CRP), and tumor necrosis factor alpha (TNF-α).^5^ These responses contribute to endothelial dysfunction, vascular stiffness, and metabolic dysregulation that collectively heighten risk for hypertension and insulin resistance.^6^ Experimental sleep restriction studies have demonstrated acute elevations in both systolic and diastolic blood pressure (SBP and DBP), while epidemiologic data show that chronic sleep disturbance predicts incident hypertension and type 2 diabetes.^7^ Despite these findings, sleep is an underexplored factor in cardiometabolic health,^8^ particularly in rural and underserved populations where health inequities and limited access to care exacerbate cardiometabolic burden.

Depressive symptoms share overlapping physiological pathways with sleep-related impairment, including HPA axis dysregulation and low-grade systemic inflammation.^9^ Depression has been associated with altered cortisol regulation, increased pro-inflammatory cytokine activity, and impaired glucose tolerance.^9^ These mechanisms mirror those observed in sleep-related metabolic dysfunction. The comorbidity of sleep-related impairment and depressive symptoms may therefore represent a synergistic threat to cardiovascular health, amplifying risk through convergent neurobiological, metabolic, and behavioral mechanisms.^9^ Individuals experiencing both sleep-related impairment and depressive symptoms may be particularly vulnerable to alterations in glucose regulation and BP control, even before overt disease develops.^10^

Furthermore, although cardiometabolic disease is typically associated with middle and older age, emerging evidence indicates that metabolic dysfunction and cardiovascular outcomes such as elevated BP are increasingly observed and often under-recognized in younger adults.^11^ National estimates and cohort trajectories show substantial prevalence and poor control of hypertension in early adulthood, which is concerning for downstream cardiovascular risk.^12^ Shifting behavioral patterns, psychosocial stress, and insufficient sleep in younger populations may accelerate early vascular and metabolic changes, contributing to the rising prevalence of hypertension and prediabetes in early adulthood.^3^ Understanding the extent to which sleep and depressive symptoms influence BP and glucose differently across age groups is therefore crucial for developing preventive strategies that target early, subclinical stages of disease progression.

### The Study

The aim of the present study was to assess how sleep, depressive symptoms, and modifiable indicators of cardiometabolic vulnerability influence blood pressure outcomes in a rural community setting. Given prior evidence linking both behavioral sleep and depression to cardiometabolic risk, we hypothesized that greater levels of sleep dysfunction and greater depressive symptom burden would predict elevated BP. We also explored whether BP outcomes varied by age, reflecting potential early manifestations of cardiometabolic dysregulation in younger participants.

## Methods

### Design

This study employed an exploratory descriptive cross-sectional design using survey methods and physiological screening data to assess relationships between sleep dysfunction, depressive symptoms, blood glucose, and BP.

### Setting and Sampling

An a priori power analysis using *G*Power* 3.1 indicated that a minimum of 48 participants was required to detect a moderate to large total effect (R² = .25; f² = .33; α = .05; β = .80) with up to six predictors. Using convenience sampling, we recruited *N* = 68 participants from a rural community in the southeastern United States, where cardiometabolic disease prevalence is disproportionately higher than other geographical regions.^1^ The study setting was an outdoor community festival in partnership with a local community health center. The collaboration was designed to promote health awareness and increase access to preventive screening services among local residents.

### Inclusion Criteria

Attendees aged 18 years and older who could read and understand English and who participated in on-site health screenings were invited to participate in the study.

### Variable Selection and Conceptualization

#### Explanatory Variables

Age and sex were included as nonmodifiable covariates due to their well-established biological influences on BP and metabolic regulation. BMI was included as a pragmatic anthropometric outcome indexing overall weight status, which is commonly used as a feasible proxy for adiposity and cardiometabolic burden, despite known limitations. BMI was calculated as *weight (lb)/[height (in)]² × 703*.

Sleep dysfunction and depressive symptoms were specified *a priori* as the predictors of interest in this analysis. These variables were conceptualized as modifiable psychosocial determinants influencing cardiometabolic physiology through metabolic and behavioral pathways. Sleep dysfunction was conceptualized as a composite index by combining measures of both sleep-related impairment and sleep disturbance. Sleep-related impairment and sleep disturbance assess co-occurring but distinct aspects of dysfunctional sleep. Together they provide a more complete assessment of global sleep dysfunction and align with contemporary sleep-health frameworks.^13,14^ Sleep duration was included as a behavioral indicator of sleep that is physiologically distinct from sleep-related impairment and sleep disturbance. While the composite sleep dysfunction index captured qualitative and functional dimensions of sleep dysfunction, nightly sleep hours characterized sleep quantity, a factor closely linked to metabolic, autonomic, and mood-related pathways. Including sleep duration alongside depressive symptoms and sleep dysfunction allowed us to differentiate the effects of sleep quantity versus sleep-related symptomatology on BP outcomes and to explore their potential additive or interactive contributions to early hemodynamic dysregulation.

### Instruments with Validity and Reliability

#### Patient Health Questionnaire-8

Depressive symptoms were measured using the Patient Health Questionnaire-8 (PHQ-8), a validated 8-item self-report instrument assessing depressive symptom severity over the past two weeks. Items are rated on a 4-point scale (0 = not at all to 3 = nearly every day), yielding total scores from 0 to 24, with higher scores indicating greater symptom burden. The PHQ-8 demonstrates strong internal consistency (α ≈ .86–.89), good test– retest reliability, and established criterion and construct validity in community and clinical samples.^15^

#### PROMIS Sleep-Related Impairment and Sleep Disturbance

Sleep-related impairment and sleep disturbance were assessed using the Patient-Reported Outcomes Measurement Information System (PROMIS) Sleep-Related Impairment Short Form (v1.0), which assesses perceived functional impairment due to perceived problems with sleep quality and continuity, and PROMIS Sleep Disturbance Short Form (v1.0), which captures self-reported difficulties with sleep initiation, maintenance, and perceived sleep quality, over the past seven days. Each instrument consists of 8 items rated on a 5-point Likert scale (1 = not at all to 5 = very much). Higher scores indicate greater sleep-related impairment or disturbance. Total scores range from 8 to 40, meaning each 1-point difference reflects ∼3% of the measures’ total continuum. Thus, a 5-point increase (≈16% of the scale range) is comparable to moving from low impairment or disturbance into a clinically meaningful moderate impairment or disturbance band.^13,14^ PROMIS sleep measures demonstrate strong internal consistency (Cronbach’s α typically ≥ .85), good test–retest reliability, and established construct and criterion validity across clinical and community populations, including sensitivity to clinically meaningful differences in sleep health.^13,14^

### Sleep Duration

Average nightly sleep duration was self-reported as the average number of hours slept per night during the 14 days preceding data collection. Consistent with established cardiometabolic risk thresholds, sleep duration was categorized as short sleep (≤ 6 hours) and normal sleep (≥ 7 hours), reflecting clinically meaningful thresholds associated with cardiometabolic and psychological risk.^3^

#### Outcome Variables

##### Blood Glucose

Non-fasting blood glucose served as a unique metabolic indicator reflecting early dysglycemia. Blood glucose was modeled to characterize the extent to which depressive symptoms and sleep dysfunction corresponded with possible metabolic disruption that may precede overt cardiometabolic disease manifested as alterations in BP outcomes.

##### Body Mass Index

BMI was included as a second anthropometric outcome indexing overall weight status, which is commonly used as a feasible proxy for adiposity and cardiometabolic burden, despite known limitations. Given substantial evidence linking poor sleep and psychological distress to weight dysregulation, and the downstream influence of adiposity on BP and vascular health, BMI was examined both as a direct outcome of behavioral predictors and as a physiological intermediary along cardiometabolic pathways.

##### Blood Pressure Outcomes

BP indices were treated as dynamic indicators of vascular and systemic health, each contributing distinct physiological information. SBP reflects arterial stiffness and large-artery compliance; DBP reflects peripheral vascular resistance and is particularly informative in younger participants; and mean arterial pressure (MAP) represents average arterial load and perfusion pressure across the cardiac cycle and overall cardiovascular strain, demonstrating unique predictive value for cardiovascular outcomes above and beyond SBP and DBP.^16–18^ Collectively, these indices provide a comprehensive view of BP regulation under behavioral and psychological stressors such as sleep-related impairment and depression.

### Data Collection

Data were collected using validated self-report survey measures previously discussed, brief structured sociodemographic questionnaires, and biometric screening. Height and weight were measured using calibrated portable scales and stadiometers. BP was assessed using Omron IntelliSense Professional Blood Pressure Monitors following a standardized resting protocol. Participants were seated upright for five minutes prior to measurement. While clinical guidelines recommend repeated readings for diagnostic accuracy, single readings were obtained due to constraints of the event setting. Non-fasting blood glucose was assessed via capillary finger-stick using *Roche Diagnostics Accu-Chek* point-of-care monitors. Body mass index (BMI) was calculated as *weight (lb)/[height (in)]² × 703*.

### Data Analysis

Analyses were conducted in R using *effectsize, estimatr, semTools, MBESS, performance, correlation, psych, lavaan, and lmtest* packages. Descriptive statistics and bivariate correlations were calculated for all variables. Next, robust hierarchical linear regression models were specified a priori to assess relationships between explanatory variables and BP outcomes. Predictors were entered sequentially based on theoretical and empirical considerations. All models were adjusted for age, sex, and BMI. MAP is a deterministic function of SBP and DBP, yielding highly collinear indices. Therefore, SBP, DBP, and MAP outcomes were modeled separately. MAP was designated *a priori* as the primary outcome variable of interest due to its physiological stability and sensitivity to early hemodynamic changes.

### Data Screening

Data were screened for missingness and outliers prior to analysis. Seven cases were identified as having extreme values that were not representative of the sample, had undue influence on distributional assumptions and high leverage, and were subsequently removed from the analysis. The remaining cases were included in the final analysis.

### Ethical Considerations

Institutional Review Board approval was obtained prior to conducting this study. All participants were provided comprehensive information about the study. Written informed consent was obtained prior to data collection.

## Results

### Characteristics of the Sample

Convenience sampling was used to recruit *N* = 68 participants A total of *n* = 61 were included in the analysis presented here. Participants included in the analysis were majority female, (57% female; 43% male), with a mean age of 53.4 years (*SD* = 11.1). The sample was predominantly White (67%) and Black (33%), with no participants reporting Hispanic or Latina/o heritage. Employment status varied, with 44% employed full-time, 18% unemployed, and 38% retired. Current tobacco use was reported by 8% of participants, and 23% were former users. Over half were taking antihypertensive medication (54%), while 25% reported periodically using sleep medication. More than half of participants expressed interest in participating in future sleep research. See Table 1 for demographic information and descriptive statistics.

**Table 1.** Demographic Information and Descriptive Statistics.

| Characteristics/<br>Variable | Category | *n* | (%) | *M* | *SD* | Min/Max |
| --- | --- | --- | --- | --- | --- | --- |
| Sex | Female | 35 | 57.4 |  |  |  |
|  | Male | 26 | 42.6 |  |  |  |
| Age |  | 61 | 100.0 | 53.44 | 11.14 | 29/70 |
| Race<sup>a</sup> | Black | 20 | 32.8 |  |  |  |
|  | White | 41 | 67.2 |  |  |  |
| Employment | Full-time | 27 | 44.3 |  |  |  |
|  | Part-time | 0 | 0.0 |  |  |  |
|  | Unemployed | 11 | 18.0 |  |  |  |
|  | Retired | 23 | 37.7 |  |  |  |
| Tobacco use | No | 42 | 68.9 |  |  |  |
|  | Yes | 5 | 8.2 |  |  |  |
|  | Former | 14 | 23.0 |  |  |  |
| Medication | Sleep | 15 | 24.6 |  |  |  |
|  | Diabetes | 10 | 16.4 |  |  |  |
|  | Hypertension | 33 | 54.1 |  |  |  |
| Blood glucose | Metabolic | 61 | 100.0 | 130.0 | 71.9 | 70/298 |
| Body mass index | Metabolic | 61 | 100.0 | 34.0 | 6.5 | 21/50 |
| Depressive symptoms | Behavioral | 61 | 100.0 | 4.8 | 4.13 | 0/19 |
| Sleep disturbance | Behavioral | 61 | 100.0 | 57.8 | 9.0 | 40.8/70.9 |
| Sleep-related impairment | Behavioral | 61 | 100.0 | 49.2 | 53.7 | 30.1/68.3 |
| Sleep duration (hours) | Behavioral | 61 | 100.0 | 6.13 | 1.60 | 3/10 |
| Systolic blood pressure | Cardiovascular | 61 | 100.0 | 149.0 | 19.6 | 108/191 |
| **Diastolic blood pressure** | Cardiovascular | 61 | 100.0 | 93.8 | 11.9 | 74/118 |
| **Mean arterial pressure** | Cardiovascular | 61 | 100.0 | 112.0 | 12.8 | 86/139 |
| *Note.* $N = 61$. <sup>a</sup> All participants reported being non-Hispanic. | | | | | | |

### Correlational Analysis

Age demonstrated inverse correlations with DBP (*r* = -.55, *p* < .001), and MAP (*r* = - .55, *p* < .001). Sex was positively related to sleep duration (*r* = .779, *p* < .001). Blood glucose was positively associated with BMI (*r* = .41, *p* = .028), sleep dysfunction (*r* = .53, *p* < .001),), SBP (*r* = .59, *p* < .001), and MAP (*r* = .48, *p* = .003). Depressive symptoms were positively associated with sleep dysfunction (*r* = .62, *p* < .001), DBP (*r* = .607, *p* < .001), and MAP (*r* = .526, *p* < .001). Depressive symptoms were negatively related to sleep duration (*r* = -.43, *p* = .015). Complete bivariate correlations are shown in Table 2.

**Table 2.** Bivariate Correlations.

| Variable | Age | Sex | BG | BMI | PHQ8 | SDYS | SDUR | SBP | DBP | MAP |
| --- | --- | --- | --- | --- | --- | --- | --- | --- | --- | --- |
| Age |  |  |  |  |  |  |  |  |  |  |
| Sex | -.083 |  |  |  |  |  |  |  |  |  |
| BG | -.177 | -.445* |  |  |  |  |  |  |  |  |
| BMI | -.318 | -.352 | .410* |  |  |  |  |  |  |  |
| PHQ8 | -.282 | .108 | .005 | -.220 |  |  |  |  |  |  |
| SDYS | -.261 | .210 | .533*** | -.097 | .617*** |  |  |  |  |  |
| SDUR | .083 | -.779*** | .038 | .268 | -.432* | -.433* |  |  |  |  |
| SBP | -.387 | -.429* | .593*** | .321 | .294 | .284 | -.127 |  |  |  |
| DBP | -.554*** | .009 | .281 | .071 | .607*** | .533*** | -.364 | .554*** |  |  |
| MAP | -.546*** | -.212 | .479** | .209 | .526*** | .478** | -.293 | .854*** | .906*** |  |

*Note.* $N = 61$ . \* $p < .05$ , \*\* $p < .01$ , \*\*\* $p < .001$ . BG = Blood glucose; BMI = Body mass index; PHQ8 = Patient Health Questionnaire-8 (Depressive symptoms); SDYS = Sleep disturbance composite; SDUR = Sleep duration (hours); SBP = Systolic blood pressure; DBP = Diastolic blood pressure; MAP = Mean arterial pressure.

### Hierarchical Regression

#### Blood Glucose

Fully adjusted models (see Table 3) explained 83% of the variance in blood glucose and 33% in BMI. sleep dysfunction was the strongest predictors of blood glucose (*sr*^2^ = .51), followed by sex (*sr*^2^ = .33) and depressive symptoms (*sr*^2^ = .22). Men’s blood glucose averaged 102 mg/dl lower than women, and each 1-point increase in depressive symptoms was associated with an approximately 11-point decrease in blood glucose, and each 1-point increase in sleep dysfunction was associated with an approximately 68 mg/dl increase in blood glucose. Conversely, participants who reported sleeping ≥ 7 hours each night had blood glucose values that were about 32 mg/dl lower than those who reported sleeping ≤ 6 hours.

**Table 3.** Hierarchical Regression Predicting Blood Glucose and Body Mass Index.

| Step | Predictor | B | SE | 95% CI [B] | p | sr<sup>2</sup> | R<sup>2</sup> | aR<sup>2</sup> | ΔR<sup>2</sup> | 90% CI | AIC |
| --- | --- | --- | --- | --- | --- | --- | --- | --- | --- | --- | --- |
| **Blood Glucose** |  |  |  |  |  |  |  |  |  |  |  |
| 1 | Age | -0.97 | 0.38 | [-1.73, -0.22] | .012 | .02 | .24 | .22 |  | [0.08, 0.37] | 685 |
|  | Sex | -102.38 | 9.86 | [-122.14, -82.62] | <.001 | .33 |  |  |  |  |  |
| 2 | PHQ8 | -11.05 | 1.30 | [-13.65, -8.45] | <.001 | .22 | .24 | .20 | .0001 | [0.07, 0.36] | 685 |
| 3 | SDYS | 67.71 | 5.24 | [57.21, 78.21] | <.001 | .51 | .81 | .79 | .564 | [0.71, 0.84] | 605 |
| 4 | SDUR | -31.52 | 10.97 | [-53.51, -9.54] | .006 | .03 | .83 | .82 | .025 | [0.74, 0.86] | 598 |
| **Body Mass Index** |  |  |  |  |  |  |  |  |  |  |  |
| 1 | Age | -0.25 | 0.07 | [-0.38, -0.11] | <.001 | .16 | .25 | .22 |  | [0.17, 0.47] | 391 |
|  | Sex | -5.01 | 1.58 | [-8.56, -1.46] | .007 | .10 |  |  |  |  |  |
| 2 | PHQ8 | -0.57 | 0.23 | [-1.04, -0.10] | .018 | .07 | .33 | .29 | .084 | [0.14, 0.44] | 386 |
| 3 | SDYS | 0.52 | 0.94 | [-1.37, 2.41] | .582 | <.001 | .33 | .29 | .004 | [0.13, 0.44] | 387 |
| 4 | SDUR | -0.49 | 1.97 | [-4.44, 3.47] | .800 | <.001 | .33 | .27 | .001 | [0.12, 0.43] | 389 |

Note. Note. *N* = 61. PHQ8 = Patient Health Questionnaire-8 (Depressive symptoms); SDYS = Sleep disturbance composite; SDUR = Sleep duration (hours)

#### Body Mass Index

Fully adjusted models (see Table 3) BMI explained 34% of the variance. Age (sr^2^ = .16) and sex (*sr*^2^ = .10) were the strongest predictors (*R*^2^ = .25), with each additional year of age predicting 0.25 kg/m^2^ lower BMI, and men exhibiting BMI values that were approximately 5 kg/m^2^ lower than women. Depressive symptoms explained an additional 7% of the variance (*p* = .018). Each 1-point increase on the PHQ-8 predicted a 0.57 kg/m² lower BMI. Neither sleep dysfunction nor sleep duration explained additional variance in the model.

#### Blood Pressure Outcomes

Fully adjusted models explained 57%, 60%, and 66% of the variance in SBP, DBP, and MAP, respectively (see Tables 4-6). Sex explained the largest portion of the variance in SBP (*sr*^2^ = .29) and MAP (*sr*^2^ = .15). Compared to women, men had SBPs, DBPs, and MAPs that were approximately 30 mmHg, 6 mmHg, and 14 mmHg lower, respectively. Age consistently demonstrated inverse relationships with all BP indices, uniquely explaining 8%, 13%, and 14% of the variance in SBP, DBP, and MAP, respectively. Each additional year of age corresponded to 0.59 mmHg lower SBP, 0.45 mmHg lower DBP, and 0.50 mmHg lower MAP, indicating that younger participants exhibited higher blood pressures across all measures. BMI failed to reach significance in all BP outcome models. Sleep duration was added in the final step and was significant across BP outcomes. Participants who reported sleeping ≤ 6 hours had higher BP readings across outcomes (SBP, *B =* -18.16, *p* = .002 ; DBP, *B =* -6.30, *p* = .031; MAP, *B =* -10.33, *p* < .001). Specifically, ≤ 6 hours of sleep was associated with 18 mm Hg increase in SBP, 6 mmHg increase in DBP, and 10 mmHg increase in MAP.

**Table 4.**
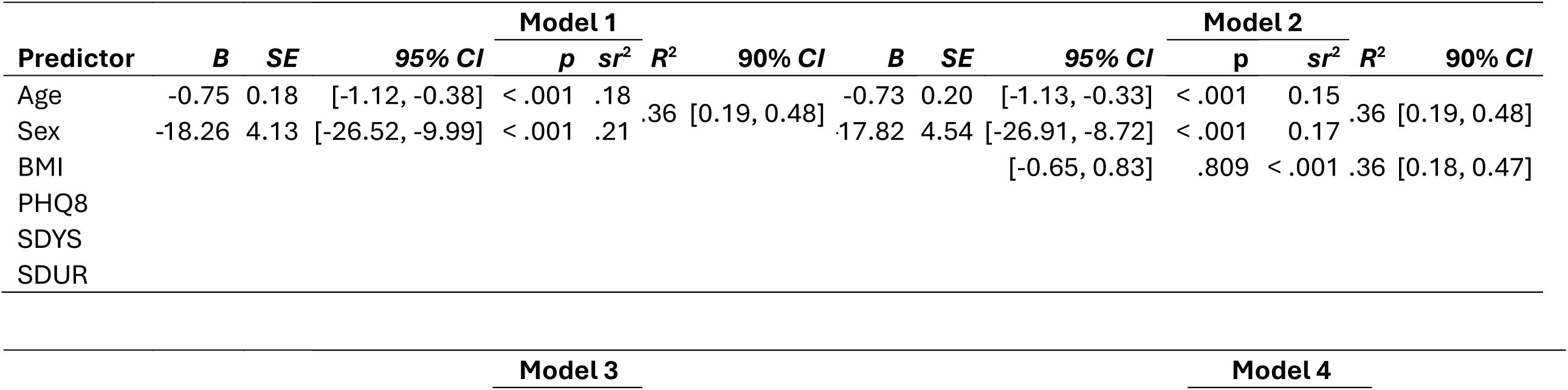
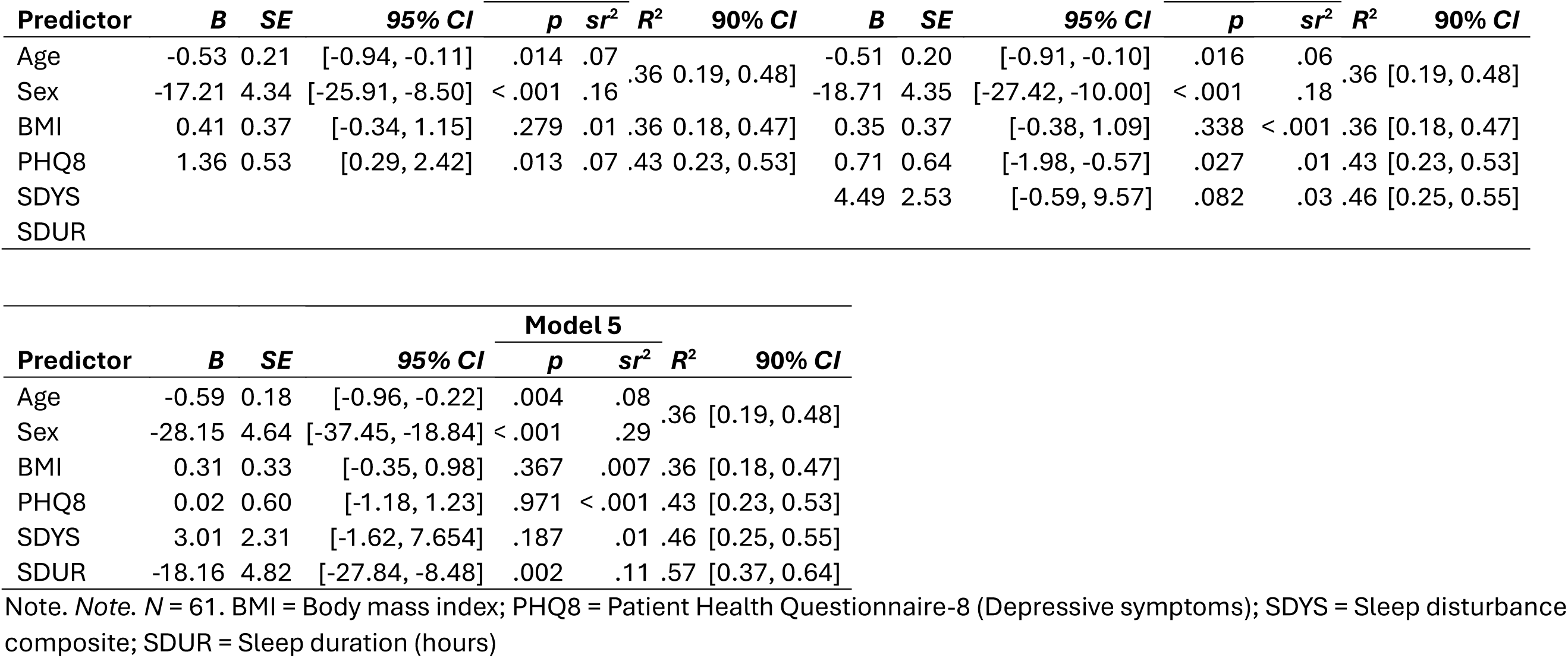
Hierarchical Regression Predicting Systolic Blood Pressure.

**Table 5.**
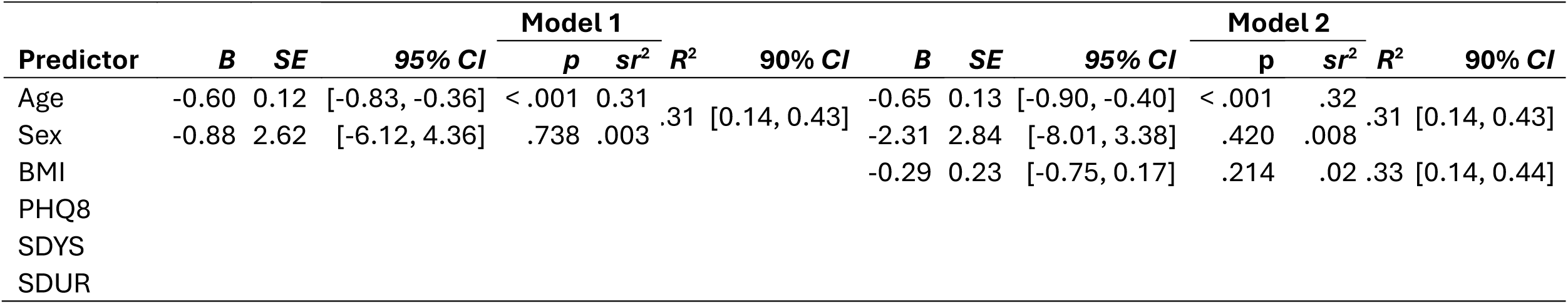
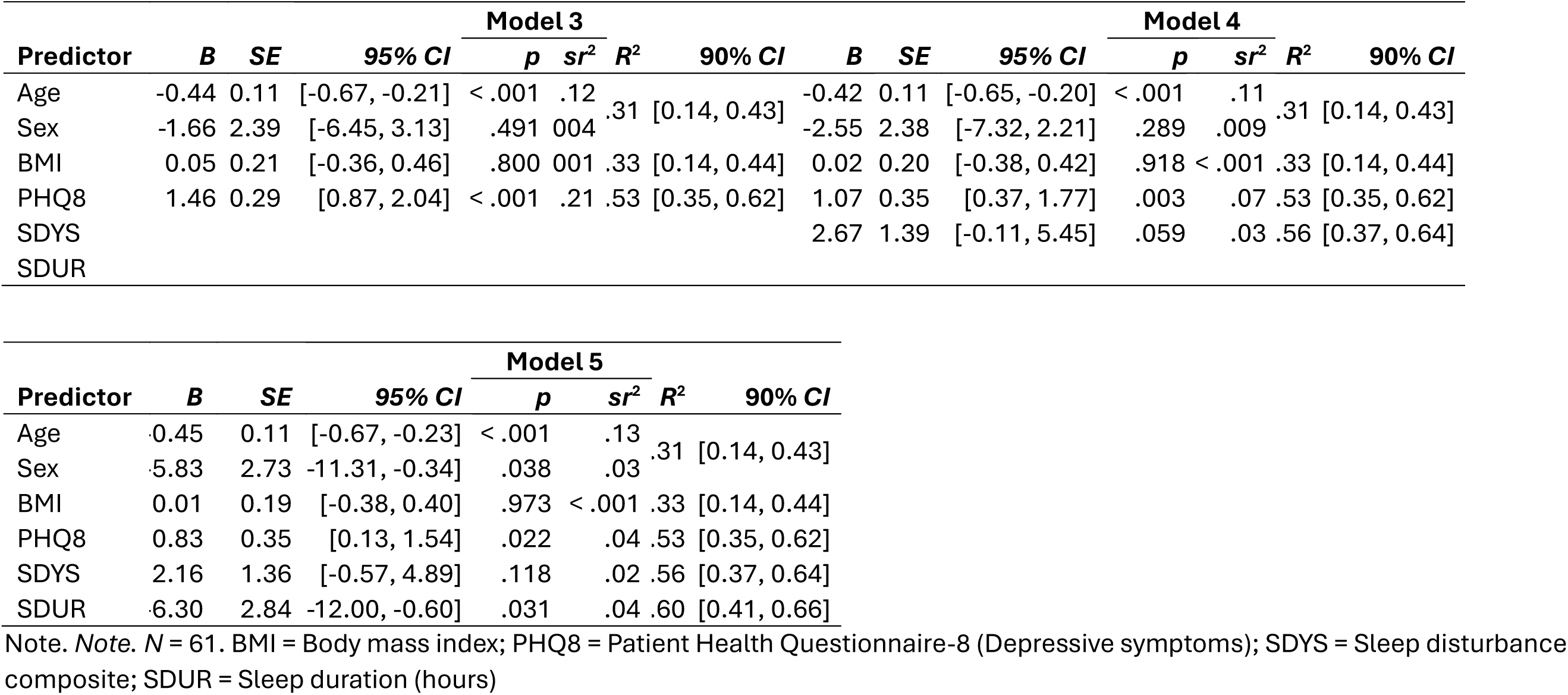
Hierarchical Regression Predicting Diastolic Blood Pressure.

**Table 6.**
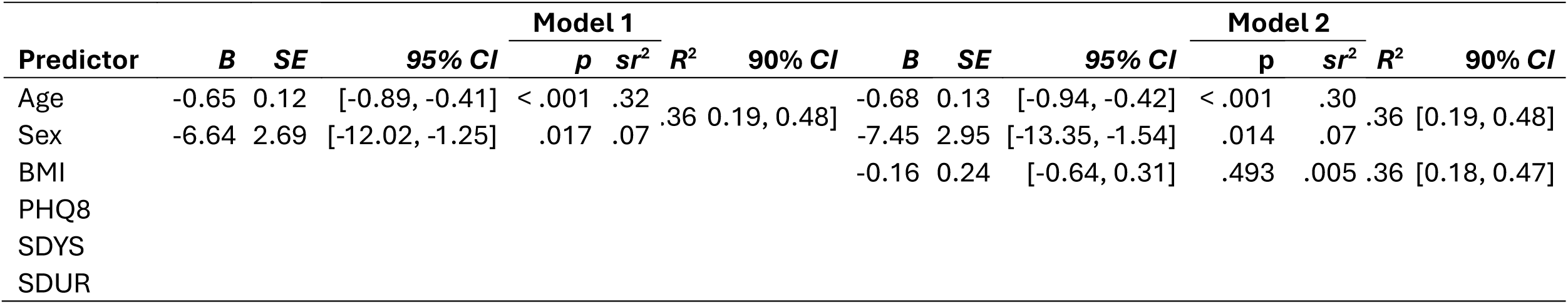
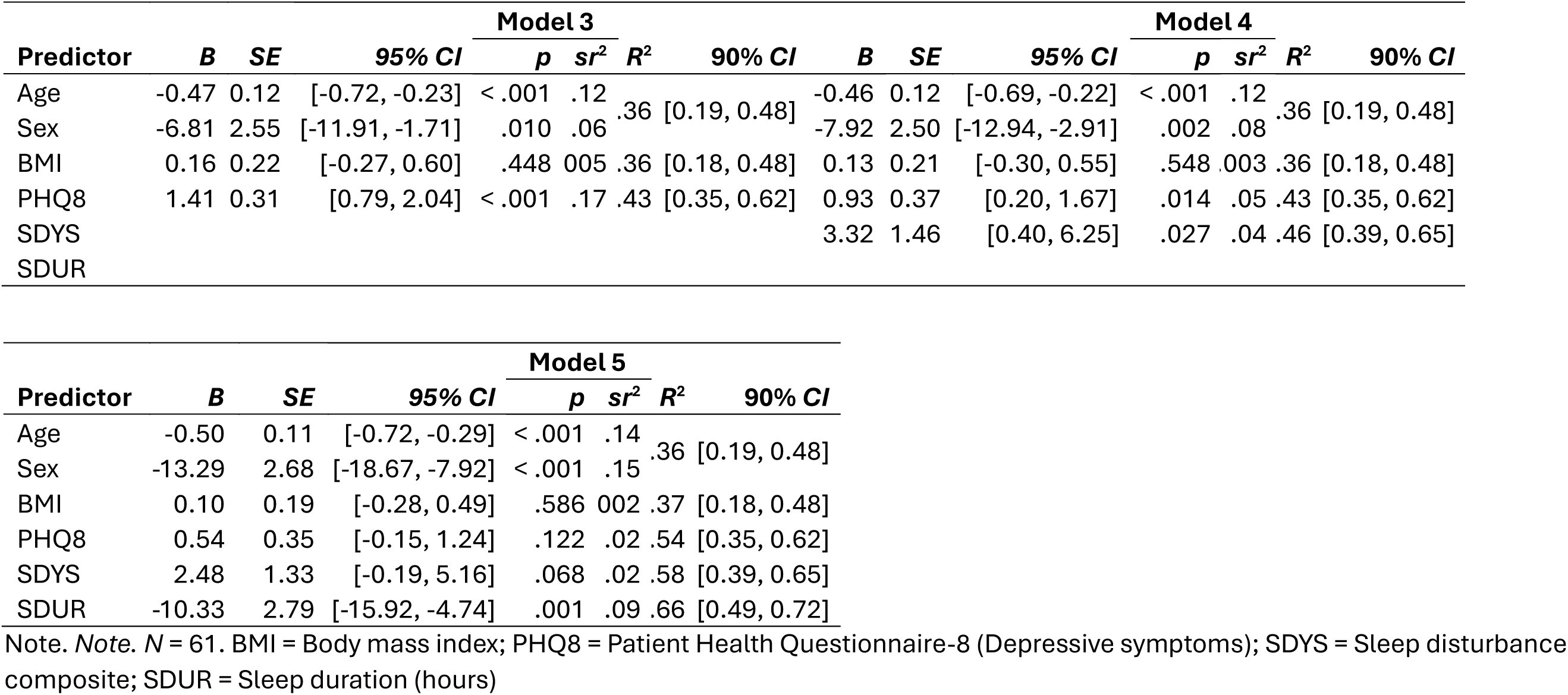
Hierarchical Regression Predicting Mean Arterial Pressure.

Depressive symptoms were associated with all BP outcomes in steps 3 (*B* = 1.36-1.46, *p* < .001 - .013) and 4 (*B* = 0.71-1.07, *p=*< .003 - .027) with each 1-point increase in depressive symptoms being associated with 1-1.5 mmHg increase BP outcomes. However, once models were adjusted for sleep dysfunction and duration, only the association between depressive symptoms and DBP was retained. Each 1-point increase in depressive symptoms was associated with an approximately 1mmHg higher DBP. The relationships between depressive symptoms and SBP, and MAP, were attenuated and lost significance.

### Exploratory Analysis

We conducted exploratory moderation analyses to examine whether sleep duration modified the relationship between depressive symptoms or sleep dysfunction and blood pressure outcomes. Interaction terms were not statistically significant and were associated with substantially larger standard errors, consistent with limited power to detect interaction effects in this small sample. Given the focus and sample size constraints, we interpreted these moderation models as exploratory.

### Sensitivity Analysis

Because the inverse association between age and blood pressure was unexpected, we conducted an exploratory sensitivity analysis to examine whether antihypertensive medication use explained this pattern. Participants who reported taking blood pressure medication had lower SBP (*M* = 144.30 mmHg, *M* age = 58.39) compared with those not taking medication (*M* = 155.61 mmHg, *M* age = 46.42), whereas group differences were negligible for DBP (*M* = 92.36 vs. *M* = 95.57 mmHg) and MAP (*M* = 109.61 vs. *M* = 115.50 mmHg).

To determine whether medication use accounted for the inverse age–BP association, we fit models including both age and medication status. Age remained a significant negative predictor of SBP (*B* = –0.57, *p* = .023), DBP (*B* = –0.68, *p* < .001), and MAP (*B* = –0.65, *p* < .001), while antihypertensive medication use showed no significant association with SBP (*B* = –5.19, *p* = .337), DBP (*B* = 4.17, *p* = .158), or MAP (*B* = 1.12, *p* = .726).

Patterns persisted in fully adjusted models that included all covariates from the primary analyses (age, sex, BMI, depressive symptoms, sleep dysfunction, and sleep duration). Age continued to predict lower SBP (*B* = –0.46, *p* = .029), lower DBP (*B* = –0.56, *p* < .001), and lower MAP (*B* = –0.54, *p* < .001). Antihypertensive medication remained non-significant for SBP (B = 5.76, *p* = .171) and MAP (B = 1.74, *p* = .527). Medication was associated with DBP (B = 5.76, *p* = .036) but again did not diminish the strength or significance of the age coefficients.

Exploratory subgroup analyses isolated the eight youngest non-medicated participants (ages 29–33), revealing markedly elevated blood pressure compared with all other non-medicated adults. The youngest subgroup had higher SBP (*M* = 169.13 mmHg, *SD* = 8.15) than all other non-medicated participants (*M* = 150.20 mmHg, *SD* = 10.78), *t*(17.13) = −5.04, *p* < .001, *d* = −1.87. Differences were greater for DBP, with the youngest adults averaging 115.88 mmHg (*SD* = 3.14) versus 87.45 mmHg (*SD* = 6.83) among older non-medicated adults, *t*(25.26) = −15.06, *p* < .001, *d* = −4.69. A similar pattern was observed for MAP (youngest: *M* = 133.63 mmHg, *SD* = 3.74; older: *M* = 108.25 mmHg, *SD* = 7.06), *t*(23.56) = −12.32, *p* < .001, *d* = −4.00. Wilcoxon tests (*p* < .001) confirmed all parametric analyses. Based on AHA MAP categories, all 8 of the youngest participants met criteria for Stage 2 hypertension, whereas all other non-medicated subgroup showed BP distributions classified in stage 1 (*M* = 102.33 mmHg, *n* = 9) and stage 2 (*M* = 113.09 mmHg, *n* = 11) range based on a single outdoor screening event, Fisher’s exact test *p* = .029.

## Discussion

This study examined how depressive symptoms and sleep relate to metabolic indicators and BP in adults attending a nurse-led screening event in a rural community. Overall, sleep dysfunction and sleep duration showed consistent associations with higher blood glucose and higher BMI. The impact of sleep dysfunction and short versus normal sleep duration was considerable, highlighting the role of sleep as a metabolic regulator and an actionable and modifiable target for early cardiometabolic intervention, given its strong links to metabolic indicators in this small rural community sample. Age and sex consistently predicted metabolic and blood pressure outcomes.

Blood pressure patterns in this sample further underscore the possibility that cardiometabolic vulnerability may occur at increasingly earlier ages. One of the most notable findings was younger participants tended to have higher BP values than older adults. This inverse age–BP pattern persisted after accounting for antihypertensive use. Mean BP values among participants not taking antihypertensive medications were elevated relative to guideline thresholds, suggesting that a subset of community adults in this setting may already be experiencing appreciable cardiometabolic strain. One possible explanation for this pattern was that older participants were more likely to be on antihypertensive medications. However, sensitivity analyses indicated that antihypertensive use did not account for the inverse age-BP associations, and the pattern of higher BP among younger participants persisted in fully adjusted models. Among participants not taking antihypertensive medications, BP values were elevated relative to current guideline thresholds, suggesting that current healthcare access or individual behaviors are missing in this at-risk group.

Exploratory subgroup analyses further highlighted findings. The eight youngest non-medicated participants (ages 29–33) exhibited markedly elevated and untreated blood pressure, with substantially higher SBP, DBP, and MAP than all other older non-medicated adults, with all eight meeting AHA criteria for Stage 2 hypertension.^19^ Although these exploratory findings are based on a small subgroup and should be interpreted with caution, the magnitude and consistency of the elevations align with national data showing a shift toward earlier onset of cardiometabolic risk. Rather than contradicting established age-related hypertension patterns, these results underscore the possibility that clinically meaningful blood pressure strain may already be present by early adulthood, particularly in rural communities with limited preventive care access. As such, our findings support growing concerns regarding early cardiometabolic vulnerability and strengthen the rationale for future research focused on younger populations.

Sleep dysfunction was positively associated with higher blood glucose, even after accounting for known physiologic drivers, age and sex. These findings align with growing evidence that the functional consequences of poor sleep such as daytime fatigue, cognitive slowing, and reduced motivation may exert more direct effects on behavior and physiology than nocturnal symptoms alone.^20^ Sleep dysfunction may promote irregular eating patterns, decreased physical activity, and heightened sympathetic activity,^21^ all of which contribute to glucose dysregulation and adiposity.^22^ The current findings are consistent with viewing sleep dysfunction as a potential marker to consider in cardiometabolic risk screening, particularly in similar rural community screening contexts, although larger studies are needed to establish clinical utility.

Depressive symptoms showed a complex profile, being linked to both blood glucose and BMI, but more closely tied to vascular outcomes. Depressive symptoms were associated with higher SBP and MAP in early steps 3 and 4, higher DBP in a fully adjusted model. These patterns are consistent with mechanistic models linking depression to heightened sympathetic arousal, endothelial dysfunction, and impaired baroreflex sensitivity,^23–25^ which may disproportionately influence DBP and MAP. However, once sleep dysfunction and duration were added in the fully adjusted models, associations between depressive symptoms and SBP and MAP lost significance, indicating that the relationship between depressive symptoms and blood pressure was attenuated by the effect of poor sleep.

Similarly, sleep dysfunction was significant across models, but its effect lost significance when models were adjusted for sleep duration, suggesting that part of the relationship between sleep dysfunction and BP may operate through sleep duration. This pattern is consistent with the possibility that insufficient sleep may overshadow the physiological effect of subjective sleep dysfunction, although formal conditional process analyses were not conducted. Our findings underscore the impact of sleep as a central biobehavioral process that shares regulatory pathways with depression, particularly autonomic activation, inflammatory signaling, and hypothalamic–pituitary–adrenal (HPA) axis dysregulation. In this context, sleep may function as both an independent contributor to vascular strain and a proximal expression of the similar physiological burden linked to depressive symptoms.

A broader point of discussion is that the pattern identified in our study is consistent with emerging concerns about cardiometabolic risk in younger adults. While hypertension is typically associated with later stages of life, a concern for older adults, findings from our study are consistent with a growing body of evidence that suggests early adulthood is a critical period during which insulin insensitivity, excess adiposity, dysglycemia, and elevated blood pressure are increasingly common but often clinically silent.^26,27^

For example, the Coronary Artery Risk Development in Young Adults (CARDIA) study, a 25-year longitudinal trial beginning at a mean baseline age of 25, demonstrated that individuals with higher blood pressure trajectories in young adulthood exhibited greater coronary artery calcification by midlife.^28^ In other studies, elevated DBP and MAP in younger adulthood has been linked to later-life cardiovascular events and target-organ damage.^29^ More recent literature suggests that youth with metabolic syndrome or persistent hyperglycemia are two to three times more likely to develop type 2 diabetes, increased carotid intima–media thickness, and structural cardiac changes in adulthood.^26,30,31^

Roughly one in five U.S. adults aged 18–39 meet criteria for hypertension, yet many remain undiagnosed or have uncontrolled blood pressure even when they have a regular source of care.^32^ At the same time, young adults are least likely to attend preventive visits and have lower rates of routine care, leading to long gaps in contact with clinicians.^33^ Furthermore, awareness of cardiovascular risk factors among young adults with hypertension, hyperlipidemia, or diabetes reflects lower perceived personal risk.^34^ These patterns suggest that younger adults may perceive their personal risk as low, seek care infrequently, and have their cardiometabolic risk underrecognized, allowing brewing metabolic dysfunction and early cardiometabolic disease to go undetected unless intentionally and systematically screened in this population. Together, these studies indicate that subclinical cardiometabolic alterations emerging in adolescence and young adulthood may go undetected, can have lasting implications for cardiovascular disease development later in life, and underscore the value of identifying early behavioral and physiological risk markers before overt hypertension or clinical cardiovascular disease emerges, especially in younger populations.

Finally, over half of participants indicated they would be interested in taking part in future sleep-related research, possibly reflecting the perceived relevance of sleep health and feasibility of conducting sleep-focused studies in this rural community. This level of interest is particularly meaningful given the well-documented barriers to research participation in rural populations, including limited access to health services, transportation constraints, and lower exposure to research opportunities.^1^ The expressed interest in sleep research suggests that community members recognize the importance of sleep for their overall health and may be motivated to participate in interventions or longitudinal studies aimed at improving sleep and related cardiometabolic outcomes. Such interest provides a strong foundation for designing community-engaged research approaches, tailoring sleep interventions to rural contexts, and developing sustainable partnerships with local health organizations to support future studies.

Future work aimed at clarifying cardiometabolic relationships or mechanistic pathways may benefit from incorporating multi-omic, biomarker, and molecular approaches that more sensitively capture early physiological dysregulation. Although this study focused on BMI and non-fasting blood glucose as metabolic indicators, cardiometabolic dysfunction begins years before these metrics meet clinical thresholds.^35^ Integrating proteomics, metabolomics, lipidomics, and cytokine/adipokine panels could reveal upstream biological signatures linking sleep health and psychological distress to metabolic and vascular outcomes. Future work could place particular emphasis on inflammatory and metabolic signaling molecules (e.g., CRP, IL-6, homocysteine, c-peptide, leptin, adiponectin) that may mediate the relationship between behavioral symptoms and cardiometabolic outcomes. Multi-omic approaches may help identify novel phenotypes such as early inflammatory–metabolic clusters or stress-reactive vascular profiles that could contribute to refining current risk stratification and inform the development of more mechanistically grounded, precision-focused interventions.

Longitudinal designed studies with larger rural samples are needed to clarify temporal dynamics, determine whether sleep-related impairment and depressive symptoms predict subsequent biomarker changes, and evaluate whether these early signals interact with social determinants of health in ways that contribute to cardiometabolic aging. Future studies could test moderation models by age, sex, race, socioeconomic status, sleep duration, and stress exposure, as well as evaluate interactions between sleep factors and depressive symptoms to identify subgroups at greatest risk. Incorporating additional behavioral pillars of cardiometabolic health, such as physical activity, diet quality, and psychological resilience, may further inform physiologically sound models.

### Strengths

A strength of this study is its integration of behavioral, psychological, and physiological data collected in a real-world rural community setting. In contrast to controlled clinical studies, this nurse-led screening event allowed for naturalistic assessment of cardiometabolic risk among adults who may not regularly seek preventive care. The use of validated tools alongside objective BP, blood glucose, and anthropometric measurements generated a multidimensional dataset capable of illustrating early behavioral and metabolic risk patterns.

The analytic strategy allowed us to explore a theoretically grounded behavioral–metabolic–vascular model, yielding estimates of individual predictors and modeling incremental variance with statistical control. This approach may help characterize associations more clearly in small community samples and offers a potentially useful template for similar studies.

### Limitations

Several limitations should be acknowledged. The cross-sectional design precludes causal inference, and the modest sample size limits power to detect small effects and to fully test complex mediational paths potential moderation effects (e.g., by age, sex, or race). BP and non-fasting blood glucose were obtained at a single time point, and fasting status likely varied, potentially increasing measurement noise. The inverse association between depressive symptoms and BMI, while statistically evident in this sample, may not generalize and should be interpreted cautiously until replicated in larger, more diverse cohorts. Future studies with larger samples should examine potential interactions between sleep dysfunction, sleep duration, and depression.

Although BMI is widely used in epidemiologic research, it is an indirect marker of adiposity. BMI does not differentiate fat from lean mass and does not capture fat distribution, such as central versus peripheral adiposity, which may be more proximal to BP and vascular risk. In the present study, BMI should therefore be interpreted as a pragmatic proxy for overall weight status, and future work incorporating more direct measures of body composition (e.g., waist circumference, waist-to-hip ratio, or imaging-based assessments) would help clarify the specific pathways linking sleep, psychological distress, adiposity, and cardiometabolic outcomes. Despite these limitations, the present findings offer preliminary evidence that sleep-related impairment and depressive symptoms may be related to cardiometabolic vulnerability through distinct yet overlapping pathways in rural adults.

### Implications for Practice

Early identification of cardiometabolic vulnerability may require looking beyond traditional metrics such as BMI, blood glucose, and BP, which often rise only after prolonged physiological dysregulation. Findings from this study are consistent with the potential value of assessing behavioral and psychological indicators, particularly sleep dysfunction, sleep duration, and depressive symptoms, as possible early clinical signals of cardiometabolic strain, even when traditional laboratory values remain within normal or mildly elevated ranges.

Nurses are well positioned to detect these early risk phenotypes in community and primary care settings. Brief, validated tools such as the PROMIS Sleep-Related Impairment short form and the PHQ-8 can be incorporated into routine assessments to identify individuals whose behavioral symptoms may contribute to metabolic disruption or autonomic imbalance. Recognizing elevated sleep dysfunction and shortened sleep hours as potential indicators of emerging metabolic dysfunction, and depressive symptoms as a possible indicator of vascular and autonomic stress, may support earlier intervention through counseling, sleep hygiene education, behavioral activation strategies, and timely referrals, before cardiometabolic illness becomes clinically diagnosable.

The observed pattern of elevated BP in younger adults underscores the importance of proactive screening across the adult lifespan, not only among those traditionally considered high risk. Nurse-led community screening events can serve as both clinical outreach and population health surveillance efforts, allowing clinicians to identify potential early maladaptive trajectories and provide preventive education in real time. Collectively, our findings are consistent with the potential value of greater attention to early behavioral and metabolic risk patterns as signals for preventive care, in addition to traditional downstream cardiometabolic endpoints.

## Conclusion

This study suggests the value of identifying early behavioral and metabolic indicators of cardiometabolic vulnerability in rural communities. Sleep dysfunction was consistently associated with metabolic indicators, whereas depressive symptoms were more closely tied to vascular and autonomic BP patterns, particularly DBP and MAP. These patterns are consistent with the possibility that behavioral indicators may reveal physiological strain well before traditional clinical metrics become abnormal. The inverse association between age and BP further suggests that younger adults may already exhibit early cardiometabolic dysfunction influenced by behavioral, environmental, and access-to-care challenges. Incorporating behavioral symptoms into routine screening may help nurses detect emerging risk trajectories and intervene earlier, particularly in rural communities. Our findings are consistent with the potential value of more proactive cardiometabolic assessment and suggest that nurse-led community screening may be a useful platform for early detection and prevention efforts.

## Data Availability

All data produced in the present study are available upon reasonable request to the authors

## Notes

### Competing Interest Statement

The authors have declared no competing interest.

### Funding Statement

Effort for this work was supported by the National Institutes of Health under award number K23AG076977 (Butts).

### Author Declarations

Institutional Review Board approval was obtained from Barton College

## References

1. Howell CR, Zhang L, Clay OJ, et al. Social determinants of health phenotypes and cardiometabolic condition prevalence among patients in a large academic health system: latent class analysis. JMIR Public Health Surveill. 2024;10:1–17. doi:10.2196/53371

2. Demissie GD, Birungi J, Shrestha A, Haregu T, Thirunavukkarasu S, Oldenburg B. The effectiveness of lifestyle interventions in reducing cardiovascular risk and risk factors in people with prediabetes: A systematic review and meta-analysis. *Nutrition*, Metabolism and Cardiovascular Diseases. 2025;35(10):104130. doi:10.1016/j.numecd.2025.104130

3. St-Onge MP, Aggarwal B, Fernandez-Mendoza J, et al. Multidimensional sleep health: Definitions and implications for cardiometabolic health: A scientific statement from the American Heart Association. Circulation: Cardiovascular Quality and Outcomes. 2025;0(0):e000139. doi:10.1161/HCQ.0000000000000139

4. Lange T, Luebber F, Grasshoff H, Besedovsky L. The contribution of sleep to the neuroendocrine regulation of rhythms in human leukocyte traffic. Semin Immunopathol. 2022;44(2):239–254. doi:10.1007/s00281-021-00904-6

5. Ballesio A, Fiori V, Lombardo C. Effects of experimental sleep deprivation on peripheral inflammation: An updated meta-analysis of human studies. Journal of Sleep Research. Published online June 5, 2025:e70099. doi:10.1111/jsr.70099

6. Direksunthorn T. Sleep and cardiometabolic health: A narrative review of epidemiological evidence, mechanisms, and interventions. IJGM. 2025;Volume 18:5831–5843. doi:10.2147/IJGM.S563616

7. Bock JM, Vungarala S, Covassin N, Somers VK. Sleep duration and hypertension: Epidemiological evidence and underlying mechanisms. Am J Hypertens. 2021;35(1):3–11. doi:10.1093/ajh/hpab146

8. Chaput JP, Stranges S. Sleep: The silent hero in cardiometabolic health. *Nutrition*, Metabolism and Cardiovascular Diseases. Published online November 2024:103782. doi:10.1016/j.numecd.2024.10.020

9. Hassamal S. Chronic stress, neuroinflammation, and depression: an overview of pathophysiological mechanisms and emerging anti-inflammatories. Front Psychiatry. 2023;14:1130989. doi:10.3389/fpsyt.2023.1130989

10. Liu C, Ye Z, Chen L, et al. Interaction effects between sleep-related disorders and depression on hypertension among adults: a cross-sectional study. BMC Psychiatry. 2024;24:482. doi:10.1186/s12888-024-05931-9

11. American Heart Association. High blood pressure a concern for adolescents and young adults in U.S. American Heart Association. 2025. Accessed July 28, 2025. https://newsroom.heart.org/news/high-blood-pressure-a-concern-for-adolescents-and-young-adults-in-u-s

12. Xia M, An J, Fischer H, Allen NB, Xanthakis V, Zhang Y. Blood pressure trajectories during young adulthood and cardiovascular events in later life. Am J Hypertens. 2024;38(1):38–45. doi:10.1093/ajh/hpae126

13. Buysse DJ, Yu L, Moul DE, et al. Development and validation of patient-reported outcome measures for sleep disturbance and sleep-related impairments. Sleep. 2010;33(6):781–792. doi:10.1093/sleep/33.6.781

14. Yu L, Buysse DJ, Germain A, et al. Development of short forms from the PROMIS sleep disturbance and sleep-related impairment item banks. Behav Sleep Med. 2011;10(1):6–24. doi:10.1080/15402002.2012.636266

15. Kroenke K, Strine TW, Spitzer RL, Williams JBW, Berry JT, Mokdad AH. The PHQ-8 as a measure of current depression in the general population. Journal of Affective Disorders. 2009;114(1):163–173. doi:10.1016/j.jad.2008.06.026

16. Flint AC, Conell C, Ren X, et al. Effect of systolic and diastolic blood pressure on cardiovascular outcomes. N Engl J Med. 2019;381(3):243–251. doi:10.1056/NEJMoa1803180

17. McCarthy CP, Natarajan P. Systolic blood pressure and cardiovascular risk: Straightening the evidence. Hypertension. 2023;80(3):577–579. doi:10.1161/HYPERTENSIONAHA.123.20788

18. Melgarejo JD, Yang WY, Thijs L, et al. Association of fatal and nonfatal cardiovascular outcomes with 24-hour mean arterial pressure. Hypertension. 2021;77(1):39–48. doi:10.1161/HYPERTENSIONAHA.120.14929

19. Jones DW, Ferdinand KC, Taler SJ, et al. 2025 AHA/ACC/AANP/AAPA/ABC/ACCP/ACPM/AGS/AMA/ASPC/NMA/PCNA/SGIM Guideline for the Prevention, Detection, Evaluation and Management of High Blood Pressure in Adults: A Report of the American College of Cardiology/American Heart Association Joint Committee on Clinical Practice Guidelines. Hypertension. 2025;82(10):e212–e316. doi:10.1161/HYP.0000000000000249

20. Hyndych A, El-Abassi R, Mader EC. The role of sleep and the effects of sleep loss on cognitive, affective, and behavioral processes. Cureus. 2025;17(5):e84232. doi:10.7759/cureus.84232

21. Greenlund IM, Carter JR. Sympathetic neural responses to sleep disorders and insufficiencies. American Journal of Physiology-Heart and Circulatory Physiology. 2022;322(3):H337–H349. doi:10.1152/ajpheart.00590.2021

22. Antza C, Kostopoulos G, Mostafa S, Nirantharakumar K, Tahrani A. The links between sleep duration, obesity and type 2 diabetes mellitus. Journal of Endocrinology. 2022;252(2):125–141. doi:10.1530/JOE-21-0155

23. Siepmann M, Weidner K, Petrowski K, Siepmann T. Heart rate variability: A measure of cardiovascular health and possible therapeutic target in dysautonomic mental and neurological disorders. Appl Psychophysiol Biofeedback. 2022;47(4):273–287. doi:10.1007/s10484-022-09572-0

24. Valenza G. Depression as a cardiovascular disorder: Central-autonomic network, brain-heart axis, and vagal perspectives of low mood. Front Netw Physiol. 2023;3. doi:10.3389/fnetp.2023.1125495

25. Waclawovsky AJ, De Brito E, Smith L, Vancampfort D, Da Silva AMV, Schuch FB. Endothelial dysfunction in people with depressive disorders: A systematic review and meta-analysis. Journal of Psychiatric Research. 2021;141:152–159. doi:10.1016/j.jpsychires.2021.06.045

26. Agbaje AO, Zachariah JP, Barker AR, et al. Persistent hyperglycemia and insulin resistance with the risk of worsening cardiac damage in adolescents: A 7-year longitudinal study of the alspac birth cohort. Diabetes Care. 2025;48(6):896–904. doi:10.2337/dc24-2459

27. Gooding HC, Gidding SS, Moran AE, et al. Challenges and opportunities for the prevention and treatment of cardiovascular disease among young adults: Report from a national heart, lung, and blood institute working group. Journal of the American Heart Association. 2020;9(19):e016115. doi:10.1161/JAHA.120.016115

28. Allen NB, Siddique J, Wilkins JT, et al. Blood pressure trajectories in early adulthood and subclinical atherosclerosis in middle age. JAMA. 2014;311(5):490–497. doi:10.1001/jama.2013.285122

29. Yang L, Magnussen CG, Yang L, Bovet P, Xi B. Elevated blood pressure in childhood or adolescence and cardiovascular outcomes in adulthood. Hypertension. 2020;75(4):948–955. doi:10.1161/HYPERTENSIONAHA.119.14168

30. Magnussen CG, Koskinen J, Chen W, et al. Pediatric metabolic syndrome predicts adulthood metabolic syndrome, subclinical atherosclerosis, and type 2 diabetes mellitus but is no better than body mass index alone. Circulation. 2010;122(16):1604–1611. doi:10.1161/CIRCULATIONAHA.110.940809

31. Wilson DP, Shah AS. Metabolic syndrome in youth - A fresh look at an old problem. Journal of Clinical Lipidology. 2025;19(4, Supplement):4–14. doi:10.1016/j.jacl.2025.03.005

32. Ostchega Y, Fryar CD, Nwankwo T, Nguyen DT. Hypertension prevalence among adults aged 18 and over: United States, 2017–2018. 2020. Accessed November 26, 2025. https://www.cdc.gov/nchs/products/databriefs/db364.htm

33. Ilango SM, Hest R, Schmidt A, McManus MA, Call K, White PH. Gaps in care among adolescents and young adults in the U.S. Am J Prev Med. 2025;69(4):107957. doi:10.1016/j.amepre.2025.107957

34. Bucholz EM, Gooding HC, de Ferranti SD. Awareness of cardiovascular risk factors in U.S. young adults aged 18–39 years. Am J Prev Med. 2018;54(4):e67–e77. doi:10.1016/j.amepre.2018.01.022

35. Mietus-Snyder M, Perak AM, Cheng S, et al. Next generation, modifiable cardiometabolic biomarkers: Mitochondrial adaptation and metabolic resilience: A scientific statement from the American Heart Association. Circulation. 2023;148(22):1827–1845. doi:10.1161/CIR.0000000000001185

